# Genome-wide gene-diet interaction analysis in the UK Biobank identifies novel effects on Hemoglobin A1c

**DOI:** 10.1101/2020.12.23.20248650

**Authors:** Kenneth E. Westerman, Jenkai Miao, Daniel I. Chasman, Jose C. Florez, Han Chen, Alisa K. Manning, Joanne B. Cole

## Abstract

Diet is a significant modifiable risk factor for type 2 diabetes (T2D), and its effect on disease risk is under partial genetic control. Identification of specific gene-diet interactions (GDIs) influencing risk biomarkers such as glycated hemoglobin (HbA1c) is a critical step towards developing precision nutrition for T2D prevention, but progress has been slow due to limitations in sample size and accuracy of dietary exposure measurement. We leveraged the large sample size of the UK Biobank (UKB) cohort and a diverse group of dietary exposures, including 30 individual dietary traits and 8 empirical dietary patterns, to conduct genome-wide interaction studies in ∼340,000 European-ancestry participants to identify novel GDIs influencing HbA1c. We identified five variant-dietary trait pairs reaching genome-wide significance (*p* < 5×10^−8^): two involved dietary patterns (meat pattern with rs147678157 and a fruit &vegetable-based pattern with rs3010439) and three involved individual dietary traits (bread consumption with rs62218803, dried fruit consumption with rs140270534, and milk type [dairy vs. other] with 4:131148078_TAGAA_T). All of these were affected minimally by adjustment for geographical and lifestyle-related confounders, and four of the five variants lacked any genetic main effect that would have allowed their detection in a traditional genome-wide association study for HbA1c. Notably, multiple loci near transient receptor potential subfamily M genes (*TRPM2* and *TRPM3*) were identified as interacting with carbohydrate-containing food groups. Some of these interactions showed nominal replication in non-European ancestry UKB subsets, as well as association using alternative measures of glycemia (fasting glucose and follow-up HbA1c measurements). Our results highlight relevant GDIs influencing HbA1c for future investigation, while reinforcing known challenges in detecting and replicating GDIs.

## INTRODUCTION

Diet is a major modifiable risk factor for type 2 diabetes and related cardiometabolic traits but a complete understanding of the therapeutic potential of dietary intervention is limited by the high inter-individual variability in glycemic response [1]. The relationship between diet and hyperglycemia is known to be influenced by genetics [2,3], suggesting that gene-diet interaction (GDI) analysis for glycemic traits has the potential to uncover novel genetic variants that modify the effect of dietary intake on glycemic response and diabetes risk. This approach can contribute to our understanding of diet-related health disparities between populations and inform the development of precision nutrition.

Despite much interest in GDIs over the past two decades, there have been few robust discoveries [4]. This can be attributed in part to the low statistical power of gene-environment interaction (GEI) analysis in general, which requires approximately four times the sample size of a genetic main effect analysis to detect associations of similar strength [5,6]. The recent availability of both cohorts with hundreds of thousands of individuals, such as the UK Biobank (UKB), and computationally-efficient software programs for biobank-scale GEI analysis represents an opportunity to overcome power limitations and identify novel interactions of modest effect size [7,8]. Recent analyses at loci with genetic main effects in UKB have identified GEIs impacting body mass index (BMI), using a series of exposures including, but not limited to, food frequency questionnaire (FFQ)-based diet measurements [9,10]. However, the use of self-reported dietary data as environmental exposures compounds the power limitations of GEI analysis. FFQ-based intake estimates are imprecise and influenced by numerous potentially confounding socio-cultural factors [11], which may bias interaction estimates toward the null hypothesis and further hamper discovery. Empirical dietary patterns that combine correlated single foods or nutrients provide one approach to deriving more meaningful and robust representations of dietary behaviors. For example, a recent genome-wide association study of dietary behaviors in UKB identified genetic signals that were unique to either individual foods or dietary patterns, emphasizing the potential for discovery when using complementary and multivariate dietary traits [12].

Here, we leveraged the large-scale UKB dataset along with a diverse set of dietary exposures to search for novel gene-diet interactions impacting glycated hemoglobin (HbA1c), a blood biomarker of hyperglycemia and diagnostic biomarker for T2D. Primary interaction analysis was conducted genome-wide in a subset of unrelated individuals of European ancestry without diabetes (N ∼ 340,000), using a series of individual diet traits (e.g. cooked vegetable intake and level of fat in milk) as well as principal components analysis (PCA)-based dietary patterns. We identified five loci reaching genome-wide significance (p < 5×10^−8^) and conducted comprehensive characterization of their associations with sensitivity analyses incorporating known confounders, multi-ancestry replication, and analysis using related glycemic traits.

## RESULTS

### Dietary behaviors associate with HbA1c

Thirty dietary traits capturing type and overall frequency of a variety of foods and beverages were derived from brief FFQs in an unrelated subset of European-ancestry participants from the UK Biobank (workflow in Fig. 1; UKB population characteristics in Supp. Table S1). These traits included semi-quantitative estimates of consumption frequency (such as cooked vegetables, bread, coffee, and alcohol), food or drink types (such as milk type [dairy vs. non-dairy milk] and decaffeinated vs. caffeinated coffee), and avoidance of specific foods (such as wheat) (Supp. Table S2). Moderate correlation between many traits, with a maximum Pearson correlation of 0.43 between beef and lamb/mutton intake (Supp. Fig. S1) motivated the derivation of both global and food sub-group dietary patterns, combining correlated foods into summary variables using PCA. Eight empirical dietary patterns (dietary principal components, or dPCs) were derived from the first principal component of seven group-specific PCA on related foods (fruit &vegetable, drinks, fish, meat, grains, dairy, and food avoidance; see Supp. Table S2) and one PCA on the full set of 30 dietary traits (global dPC) (Fig. 2a). The global dPC, which explained 9.4% of the total variance in all dietary traits, remained reflective of a general “prudent” dietary pattern as has been previously observed in UKB [12].

**Figure 1:**
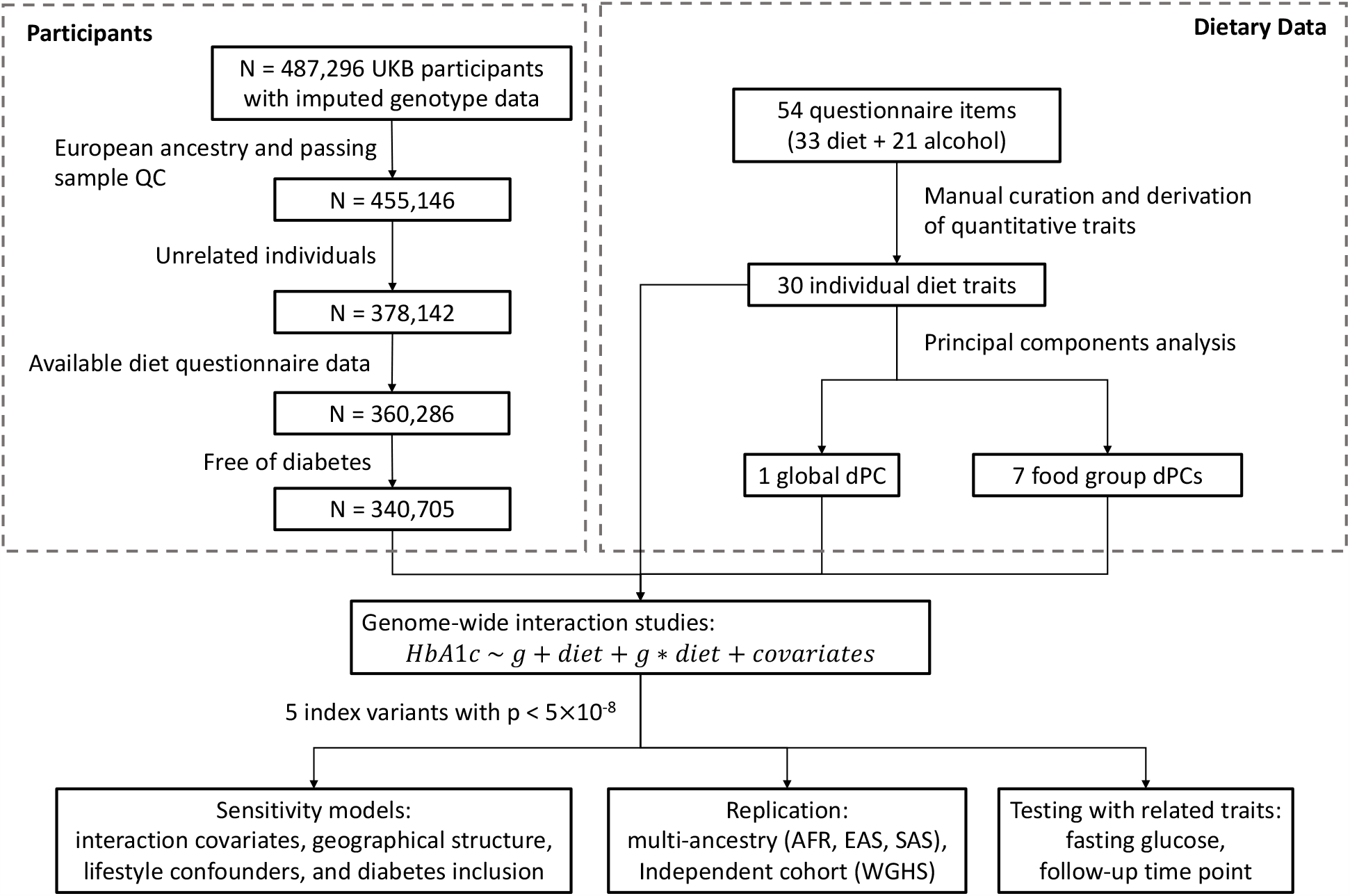
Participant selection and analysis workflow. UK Biobank participants underwent filtering based on genotyping quality control, relatedness, ancestry, availability of dietary data, and diabetes status. Dietary traits were curated from FFQ responses and dietary patterns were derived from principal components analysis. All 38 dietary traits were used in genome-wide interaction studies for HbA1c, with follow-up sensitivity, replication, and within-population analyses using related glycemic traits.

**Figure 2:**
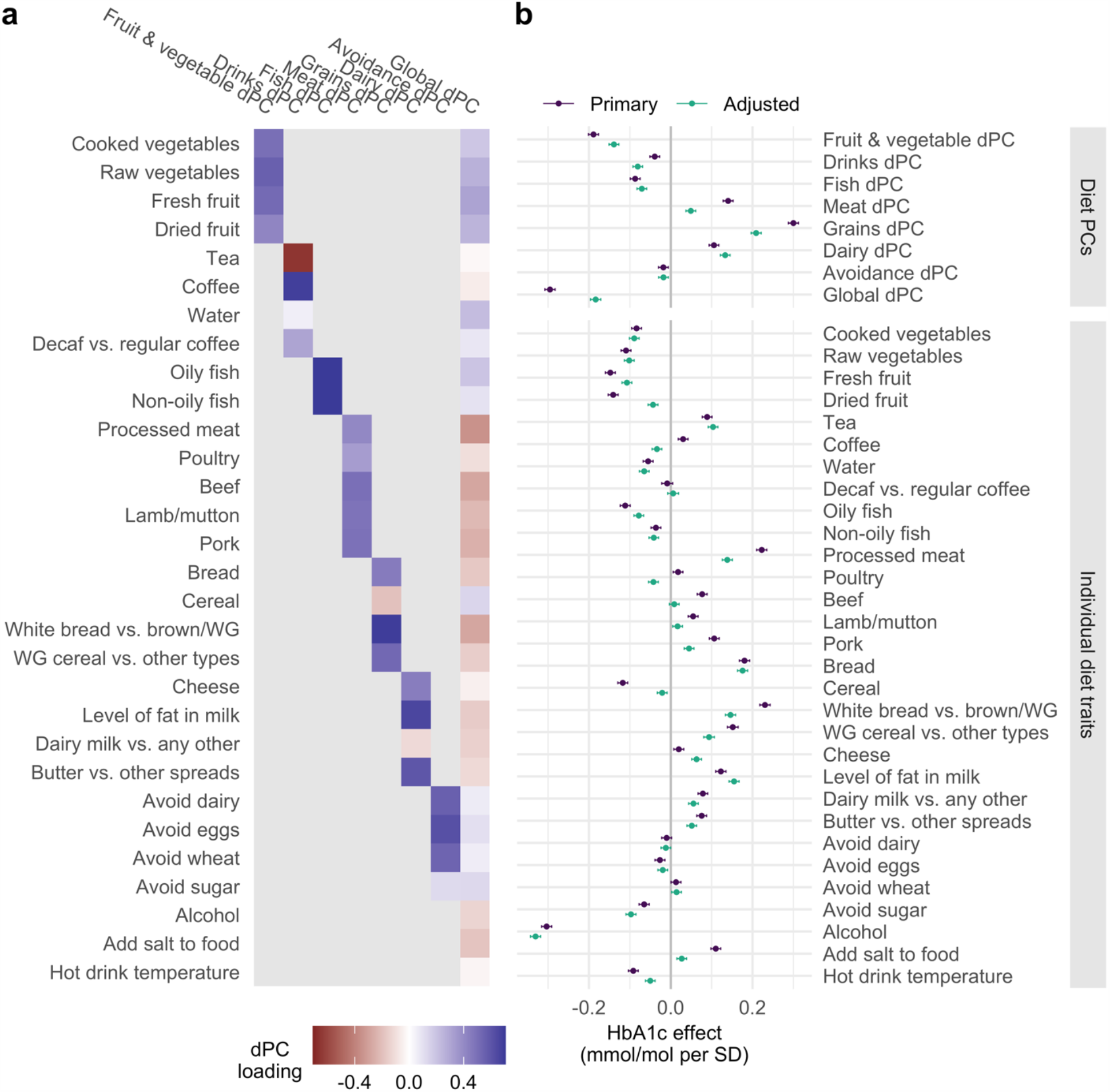
Dietary traits and associations with HbA1c. a) Heatmap displays loadings for each of the 30 dietary traits characterizing the seven food group dPCs and one global dPC. Higher positive loadings denote higher contributions of the trait (*y*-axis) to the associated dPC (*x*-axis). b) Forest plot of coefficients from regression of HbA1c on each dietary trait. Models included either the same set of covariates as in the primary GWIS (“Primary”; purple) or those from sensitivity model 3 (“Adjusted”; adjusted for birthplace, assessment center, BMI, smoking, educational attainment, and physical activity; purple). Error bars correspond to 95% confidence intervals (though negligible in size due to the scale).

To first understand the relationship between these dietary traits and HbA1c independently of genetic background, we regressed HbA1c on each of the dietary traits, finding associations at the majority of dietary patterns and individual traits (32 of 38 traits at Bonferroni-corrected p < 0.05 / 38 = 0.0013) (Fig. 2b). The strongest associations were observed for the grains dPC (0.30 mmol/mol HbA1c per standard deviation, 95% CI: 0.29-0.31, *p* < 1×10^−300^), which loads most strongly on bread type (increased white vs. brown or whole grain bread), the “prudent” global dPC (0.30 mmol/mol HbA1c per standard deviation, 95% CI: 0.28-0.31, *p* < 1×10^−300^), which is also largely influenced by bread type, and alcohol intake (−0.316 mmol/mol HbA1c per standard deviation, 95% CI: −0.32 - −0.29, *p* < 1×10^−300^). Many of these associations were substantially attenuated by adjustment for geographic and lifestyle covariates, indicating the presence of partial confounding (between 30 and 40% reduction in effect size for the grains dPC, global dPC, and processed meat intake, though these relationships remained highly significant). Others, such as alcohol intake and bread intake, remained equally strong in the adjusted model.

### Genome-wide interaction studies

Genome-wide interaction studies (GWIS) were performed for each of the 8 dPCs and 30 individual dietary trait exposures to identify loci whose effect on HbA1c is modified by dietary intake. Across the 38 GWIS, five independent loci clumped by 500 kb windows reached genome-wide significance (p < 5×10^−8^) for their interaction effect (Fig. 3; Table 1). None of these signals passed a more stringent threshold adjusted for multiple testing of 27.3 effective traits (p < 5×10^−8^ / 27.3 = 1.83×10^−9^; see Methods).

**Table 1:**
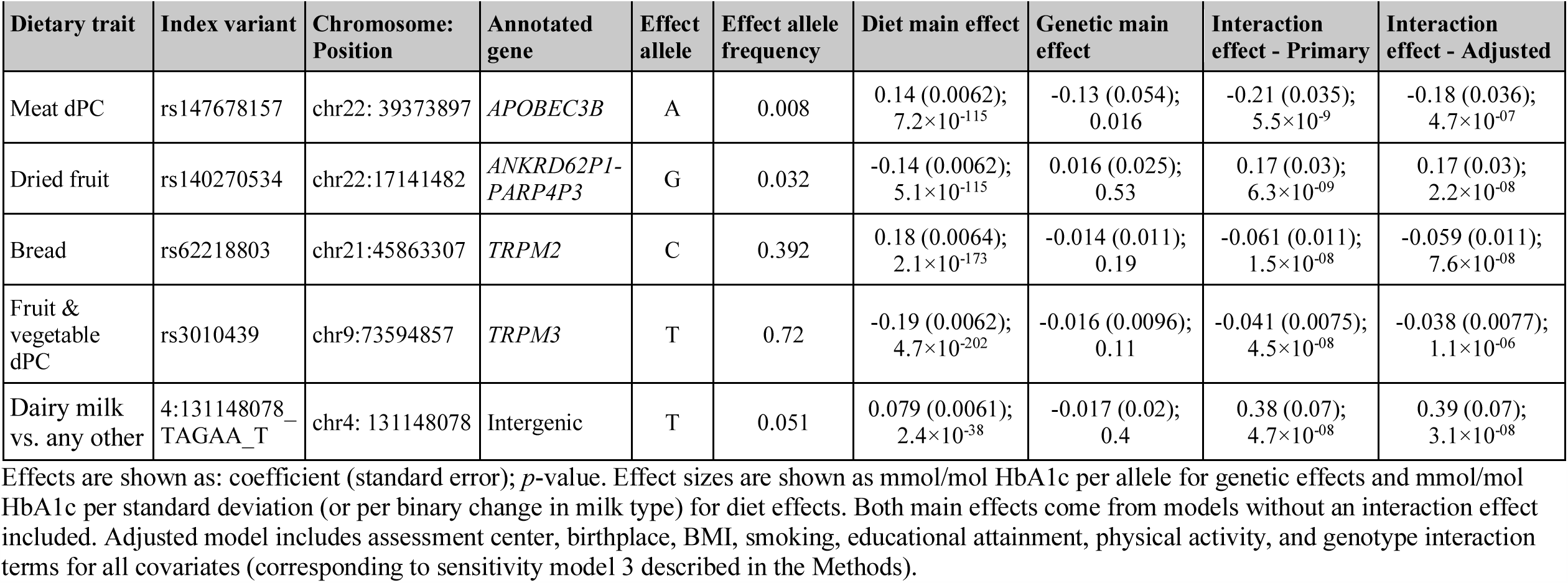
Loci reaching genome-wide significance (p < ×10-8) in primary GWIS

**Figure 3:**
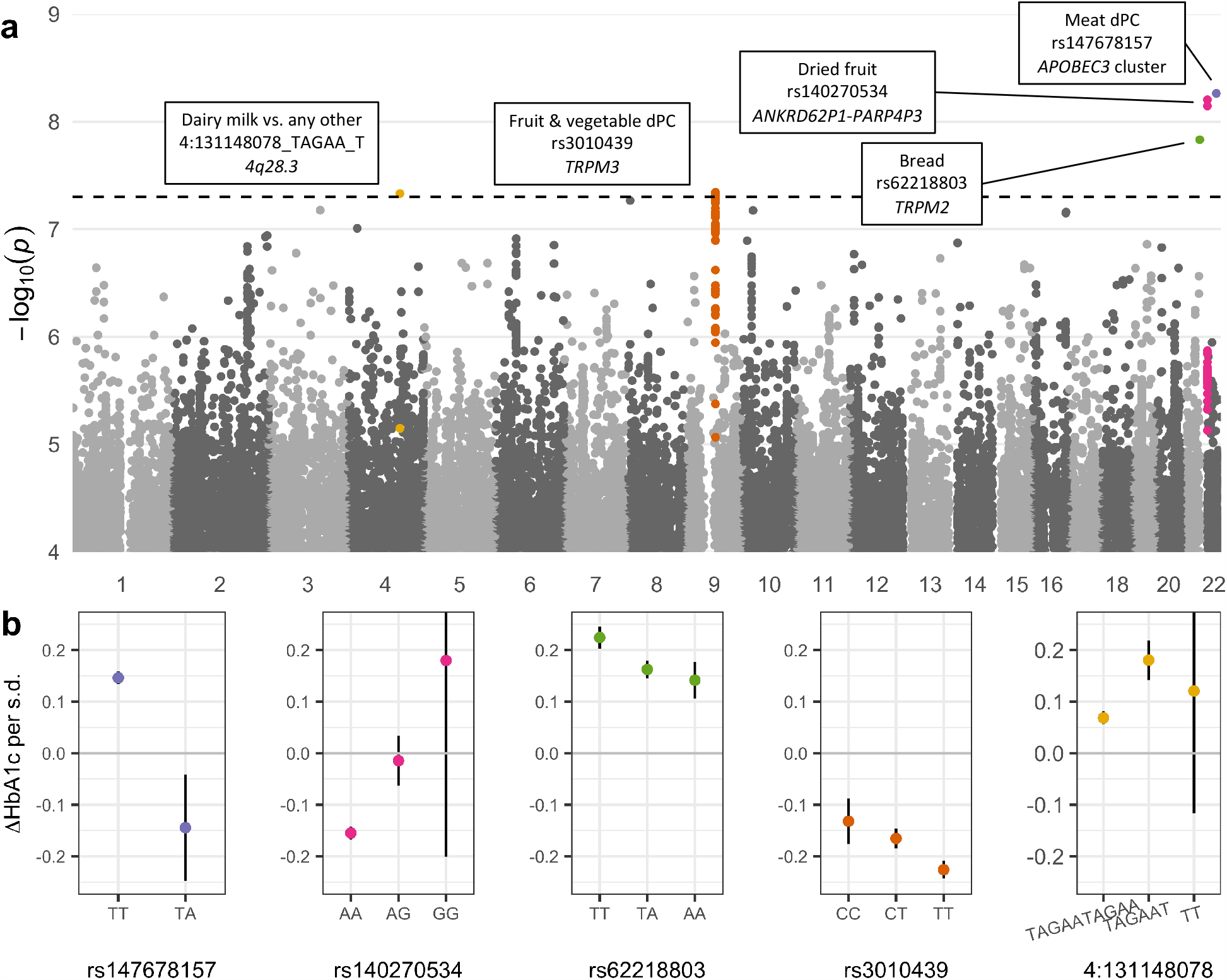
Genome-wide gene-diet interaction analysis results. a) Manhattan plot shows minimum -log_10_(*p*-values) as a function of genomic position across all 38 GWIS. Colors correspond to independent loci with index variants having *p* < 5×10^−8^, and variants are assigned to a peak if within 50kb of an index variant and having *p* < 1×10^−5^. Index variants are annotated with the trait, variant rsID (where available), and annotated gene. b) Dietary effect sizes on HbA1c (change in HbA1c associated with a standard deviation increase in the associated dietary trait; y-axis) after stratification by genotype (closest genotype based on dosages; x-axis), using the primary analysis covariates. Points and bars denote stratified main effect estimates and 95% confidence intervals from the primary model. Genotype-stratified effects are shown only for genotype groups containing >100 individuals. Effect estimates are in terms of mmol/mol HbA1c. s.d.: standard deviation.

Two of the five lead GDIs involved food group dPCs (meat dPC-rs147678157 and fruit &vegetable dPC-rs3010439) and three involved individual dietary traits (bread consumption-rs62218803, dried fruit consumption-rs140270534, and milk type [dairy vs. other]-4:131148078_TAGAA_T). Main effects of the interacting dietary traits on HbA1c were all strong, while genetic effects (as calculated from a model without any interaction term) were absent other than a small association at the meat-interacting rs147678157 (Table 1). To better characterize the interactions at our lead variants, we conducted two sets of stratified analyses: testing the genetic effects on HbA1c after stratifying our dietary exposures into quintiles and testing the dietary effects on HbA1c after stratifying by genotype. Genotypic effects all displayed a qualitative interaction pattern, in which the sign of the genetic effect differed across strata of dietary behaviors (Supp. Fig. S2). This accounts for the lack of marginal genetic effect of these variants on HbA1c. In contrast, we found evidence of quantitative interactions, in which the dietary effect changed only magnitude across genotype groups, at three out of the five loci (rs147678157, rs62218803, and rs3010439; Fig. 3b). For example, in those with two copies of the reference allele at rs62218803, each standard deviation of bread consumption associates with an HbA1c increase of 0.22 mmol/mol. The strength of this association decreases by 37% to 0.14 mmol/mol in those with two copies of the alternate allele, meaning that bread consumption may be less detrimental in these individuals.

Of the primary interactions, two were in or near transient receptor potential subfamily M genes, *TRPM2* and *TRPM3*, despite their genomic location in two different chromosomes (21 and 9, respectively). Their protein products, as well as other transient receptor potential (TRP) family proteins, have been implicated in insulin secretion and downstream glycemia [13]. Furthermore, some of these index variants had marginal interaction effects on HbA1c with other dietary traits (such as the meat dPC-interacting rs147678157 with the global dPC and tea intake, as well as the dried fruit-interacting rs140270534 with the fruit &vegetable dPC), whereas other lead GDIs were apparently unique to the dietary exposure used in discovery (such as the milk type-interacting 4q28.3 variant; Supp. Table S3). Each dPC-interacting variant showed some degree of interaction with at least one of that dPC’s constituent traits, indicating that the use of PCA-based dietary patterns increased our power for the detection of interactions. For example, *p*-values for rs147678157 were between 1×10^−6^ and 1×10^−3^ for four of five individual meat consumption traits, while the meat dPC interaction reached *p* = 5.45×10^−9^.

Sensitivity models were conducted at all five index variants to assess the potential effects of confounding and interaction model biases (see Methods). Interaction effect estimates were consistent after adjustment for genotype × covariate interactions (sensitivity model 1 [SM1]), birthplace and study center (SM2), and BMI, smoking, physical activity, and educational attainment (SM3) (Supp. Fig. S3, Supp. Table S4). The effects were only modestly attenuated when including individuals with diabetes (SM4), whose diagnosis and treatment would directly affect HbA1c via both behavioral changes and medication use. The small degree of change in the interaction effect estimates across these sensitivity models indicates that the observed relationships are not primarily due to confounding by known correlates of diet such as socioeconomic status or a generally healthy lifestyle.

### Replication across ancestries and cohorts

To assess the translation of these GDIs in non-European populations, we conducted a multi-ancestry replication of the five significant interactions in individuals of West African (AFR; N = 4,763 after sample QC), East Asian (EAS; N = 1,936), and South Asian (SAS; N = 5,849) ancestry based on the same dietary data in UKB. One variant-trait pair, rs62218803 and bread consumption (slices per day), was nominally significant (*p* = 0.022) and directionally consistent in the East Asian ancestry group. Three nominal and directionally consistent interactions were observed for dPC-interacting variants with the individual contributing dietary traits (Supp. Table S5): one for the meat dPC locus (processed meat-rs147678157 in SAS, *p* = 0.022), and two for the fruit &vegetable dPC locus (fresh fruit-rs3010439 in AFR, *p* = 0.008 and cooked vegetables-rs3010439 in SAS, *p* = 0.012). Overall, the majority of variant-trait-ancestry combinations did not show meaningful replication, though we note the limited power afforded by the much smaller sample size.

To explore this further, we gathered all variant-trait pairs reaching suggestive significance (*p* < 1×10^−5^) in the primary analysis and tested for an enrichment of shared interaction coefficient signs in the multi-ancestry analysis. As shown in Table 3, we did not find evidence of such an enrichment in any of the ancestries, an observation that could be attributed partially to lower sample sizes, differences in dietary intake and its socio-cultural correlates, and known differences in genetic architecture (e.g. haplotype blocks, allele frequencies, and causal variants).

**Table 2:** Multi-ancestry replication of index variant-trait pairs from the primary analysis

| Dietary trait | Index variant | Primary analysis |  | Replication |  |  |  |  |  |
| --- | --- | --- | --- | --- | --- | --- | --- | --- | --- |
|  |  | EUR AF | EUR Effect (P) | AFR AF | AFR Effect (P) | EAS AF | EAS Effect (P) | SAS AF | SAS Effect (P) |
| Meat dPC | rs147678157 | 0.008 | -0.207<br>($5.5 \times 10^{-9}$) | 0.001 | -1.625<br>(0.2) | 0.002 | 1.332<br>(0.72) | 0.003 | 1.471<br>(0.091) |
| Dried fruit | rs140270534 | 0.032 | 0.174<br>($6.3 \times 10^{-9}$) | 0.003 | -0.622<br>(0.62) | 0.066 | 0.119<br>(0.67) | 0.078 | -0.081<br>(0.67) |
| Bread | rs62218803 | 0.392 | -0.061<br>($1.5 \times 10^{-8}$) | 0.437 | 0.006<br>(0.96) | 0.481 | -0.377<br>(0.022) | 0.438 | -0.057<br>(0.58) |
| Fruit & vegetable dPC | rs3010439 | 0.72 | 0.041<br>($4.5 \times 10^{-8}$) | 0.568 | -0.102<br>(0.22) | 0.632 | -0.058<br>(0.59) | 0.727 | 0.117<br>(0.08) |
| Dairy milk vs. any other | 4:131148078_TAGAA_T | 0.051 | 0.382<br>($4.7 \times 10^{-8}$) | 0.006 | -1.312<br>(0.27) | 0.029 | 0.029<br>(0.97) | 0.096 | -0.36<br>(0.49) |

Effects are shown as: coefficient ( $p$ -value). Coefficients are in units of: mmol/mol HbA1c per allele per standard deviation of the dietary trait. AF: allele frequency.

**Table 3:** Aggregate replication and validation of suggestive (p < 1×10^−5^) index variant-trait pairs

| Dataset | Sample size | # variants tested<sup>1</sup> | % directionally consistent | P<sup>2</sup> |
| --- | --- | --- | --- | --- |
| West African | 4763 | 958 | 52% | 0.16 |
| East Asian | 1936 | 486 | 48% | 0.25 |
| South Asian | 5849 | 738 | 51% | 0.96 |
| Follow-up | 10644 | 959 | 66% | $8.3 \times 10^{-26}$ |
| Fasting glucose | 10879 | 959 | 57% | $3.9 \times 10^{-6}$ |

<sup>1</sup>Variant numbers vary across ancestries due to the MAF > 0.005 criterion
<sup>2</sup>Chi-square test $p$ -value for shared interaction effect signs with primary analysis

### Follow-up analyses at lead GDIs

To further explore these top loci, we tested for GDIs using follow-up FFQ and HbA1c data from a subset of UKB individuals from the primary analysis that participated in a repeat assessment center visit between 2012-2013 (N = 10,644) and for GDIs using fasting glucose from the subset of individuals who were fasting at their baseline visit (N = 10,879). The follow-up analysis can provide evidence for the stability of these interactions over time, and the fasting glucose analysis indicates whether these interactions are generalizable to other measures of hyperglycemia. Though significance levels were weak, possibly due to sample size limitations, the directions of the interaction effects were consistent with the primary analysis for all variant-trait pairs (Supp. Table S6). The strongest associations included the meat dPC-rs147678157 interaction impacting fasting glucose (*p* = 0.02) and the milk type-4:131148078_TAGAA_T locus interaction evaluated at follow-up (*p* = 0.005). Given the generally lower statistical power for these analyses, we tested for an enrichment of directional consistency compared to the primary analysis across all 959 suggestive variant-trait pairs (*p* < 1×10^−5^ in any of the 38 GWIS), and found strong evidence for enrichment using a subset of the same European individuals (66% directionally consistent with *p* = 8.3×10^−26^ for follow-up and 57% directionally consistent with *p* = 3.9×10^−6^ for fasting glucose), an enrichment we did not find among our non-European replication analyses (Table 3).

To better characterize these loci at a molecular level, we used PhenoScanner to examine cross-trait associations with DNA methylation, gene expression, metabolites, and proteins, including potential proxy variants with r^2^ > 0.8 in Europeans. Variant rs3010439 (fruit &vegetable dPC-interacting), lies within an intron of *TRPM3* and has strong associations with the methylation of nearby CpG sites within *TRPM3* (including cg20555507 [*p* = 4.42×10^−14^] and proxy variant rs3095766 with cg14165911 [*p* = 8.61×10^−286^]). The other *TRPM* variant, rs62218803 (bread-interacting), is located less than 400bp downstream of *TRPM2*, yet there were no significant (*p* < 5×10^−8^) expression-quantitative trait loci (eQTLs) in any tissues contained in the associated GTEx dataset. However, approximately 100kb upstream of rs62218803, and in complete linkage equilibrium, is a missense coding variant in *PFKL* with no interaction effect (*p* = 0.92) but a strong genetic main effect on HbA1c in our analysis (rs17850433, *p* = 7.8×10^−23^; Supp. Fig. S4). Variant rs140270534 (dried fruit-interacting) lies in an intron of the pseudogene *ANKRD62P1-PARP4P3* and acts as an eQTL for *XKR3* in whole blood (*p* = 5.52×10^−13^ in the eQTLGen dataset [14]). *XKR3* codes for a homolog of the putative membrane transporter and Kell blood group component XK, but has no clear link to glycemic regulation.

To test the robustness of these interactions in an independent cohort, replication was sought in individuals of European ancestry from the United States (US)-based Women’s Genome Health Study (N = 20,095). Though we were only able to test for 3 variant-trait pairs surpassing genotype imputation quality filters, no interactions were detected at *p* < 0.05 (Supp. Table S7). We note that this lack of replication may reflect a variety of factors including heterogeneity in dietary behaviors and measurement as well as lack of statistical power.

## DISCUSSION

We have conducted the largest gene-diet interaction study for HbA1c to-date, uncovering multiple interactions between genotype and dietary traits (both individual diet questionnaire items and empirical diet patterns) that influence HbA1c. We further characterized these loci through multi-ancestry replication and other validation analyses within UKB, and showed that these loci have the potential to have a meaningful impact on the diet-hyperglycemia relationship.

HbA1c is both a diagnostic and prognostic biomarker for T2D, reflecting cumulative blood glucose over a two to three-month period prior to measurement [13]. Therefore, it provides a long-term quantitative measure of glucose levels, making it an appealing outcome for GDI analysis with dietary exposures which similarly reflect long term usual intake from FFQs. It is important to note that HbA1c is more strongly influenced by postprandial, rather than fasting, glucose blood glucose concentrations [15]. In this light, it may not be surprising that our fasting glucose analysis in a subset of the primary population did not strongly validate our top interaction associations. Furthermore, while HbA1c is a valuable continuous measure for hyperglycemia, it is also independently influenced by red blood cell characteristics, such that its genetic influences are often classified as either glycemic or erythrocytic [15]. However, based on the dietary traits used as exposures and the candidate genes identified, it does not appear that these interactions are affecting HbA1c via red blood cell traits.

The primary analysis highlighted interactions within or near two independent *TRPM* subfamily genes. *TRPM2* and *TRPM3* are members of a larger family of 28 cation-permeable TRP channels, many of which are modulated by a diverse array of natural and dietary compounds [16]. Both *TRPM2* and *TRPM3* have been shown to affect insulin secretion by beta cells in animal models [17,18], highlighting a potential role in glycemic control. The *TRPM3* variant (rs3010439), which showed an interaction with the fruit and vegetable dPC in European-ancestry individuals and fresh fruit intake in West African-ancestry individuals, is particularly interesting in that the TRPM3 protein product is inhibited by citrus fruit metabolites, specifically naringenin and hesperetin [19]. In general, our finding of interactions between *TRPM*-annotated variants and multiple carbohydrate-heavy foods (bread and fruit) provides an avenue for further exploration.

The effect sizes of the interactions identified here may be practically meaningful towards the implementation of personalized nutrition approaches. For example, based on the stratified models, an increase of one standard deviation in bread consumption (approximately 8 slices of bread per week) associates with an HbA1c increase of 0.22 mmol/mol in reference homozygotes at rs62218803 (*TRMP2*). However, this effect size is decreased by 37% to 0.14 mmol/mol in alternate allele homozygotes. Likewise, the protective effect of the fruit and vegetable-increasing dietary pattern (0.13 mmol/mol per standard deviation in reference homozygotes) is magnified by 70% in alternate allele homozygotes at rs3010439 (*TRPM3*). However, these effect estimates should be treated with caution, as they are not only susceptible to residual confounding, but also the Winner’s Curse phenomenon, in which effects from hypothesis-free studies tend to be overestimated in discovery analyses [20].

The large sample size used in this study along with the use of data-driven dietary patterns improved our statistical power to uncover gene-diet interactions of small effect size. For example, the combination of related foods into empirical dietary patterns led to the identification of genome-wide significant interaction effects that were present with only modest significance with the individual foods themselves. However, we were not able to demonstrate replication of these interactions in the independent Women’s Genome Health Study, and showed only partial nominal replication of the interactions in the secondary within-UKB analyses. This incongruence could be due to a variety of factors, including differences in dietary variability across ancestries and countries (US vs. UK), differences in the dietary questionnaires used, and differences in the extent to which these food groups act as proxies for underlying traits (such as socioeconomic status or general healthy behaviors). The difficulty in replication of gene-diet interactions was highlighted in the null findings of a systematic replication effort for gene-macronutrient interactions influencing type 2 diabetes risk in the EPIC-InterAct study [22]. The authors emphasized rigor in reporting and replication in diverse populations as strategies to combat potential false positive findings. While our in-depth replication approach, which included diverse ancestries, independent cohorts, and alternative outcomes, was an important strength of this study, it also reinforces the difficulty in replicating GDIs, even among biologically relevant candidate genes. To address known obstacles to gene-environment interaction detection [23], we suggest that future investigations seek out populations with substantial variability in the dietary behaviors of interest and use dietary datasets with greater granularity. These more precise dietary datasets can complement the dPC-based discovery approach employed here by helping with “phenotypic fine-mapping” to pinpoint the relevant dietary components.

Additional limitations should be considered when evaluating these results. First, the UKB FFQ contains a limited set of questions on high-level food behaviors and therefore does not enable the calculation of reliable nutrient or total energy intake estimates. While the use of dietary patterns, which reflect higher level patterns of behavior, may be less influenced by total energy intake, interactions involving these patterns as well as the individual dietary traits may still be susceptible to residual confounding by total energy intake. However, we note that adjustment for both BMI and physical activity, which are highly correlated with TEI [24] had a minimal impact on the results. Second, the individual dietary traits and dPCs used here are likely reflective to some extent of confounding factors such as socioeconomic status, as has been previously observed [12], and are susceptible to reverse causation (impacts of known diabetes risk or family history on dietary behaviors). While our sensitivity analyses aimed to control for socio-cultural effects on dietary intake, it remains possible that our top interactions are actually capturing additional lifestyle exposures, which, given their major differences across ethnicities and countries, would explain our limited replication.

In summary, we have leveraged the large sample size of the UK Biobank and an empirical dietary pattern-based approach to conduct a well-powered scan for gene-diet interactions influencing HbA1c. We uncovered multiple biologically relevant genetic loci that may influence the relationship between diet and hyperglycemia. Our findings indicate the value of gene-diet interaction analysis in large-scale datasets and the use of composite dietary exposure variables, while also reinforcing the persistent difficulty in detecting and replicating interactions based on self-reported dietary questionnaires. This study can serve as a basis for future investigations into the interplay of lifestyle and genetics for type 2 diabetes prevention.

## METHODS

### UK Biobank genetic data

UKB is a large prospective cohort with both deep phenotyping and molecular data, including genome-wide genotyping, on over 500,000 individuals ages 40-69 living throughout the UK between 2006-2010 [25]. Genotyping, imputation, and initial quality control on the genetic dataset have been described previously [26]. Additionally, we removed individuals flagged for failing UKBiLEVE genotype quality control, heterozygosity or missingness outliers, individuals with putative sex chromosome aneuploidy, individuals with self-reported vs. genetically inferred sex mismatches, and individuals whom withdrew consent at the time of analysis. Furthermore, only genetic variants with minor allele frequency (MAF) >0.005 and imputation INFO score > 0.5 were included, resulting in a set of approximately 11,100,000 autosomal variants for analysis. Work was conducted on genetic data release version 3, with imputation to both Haplotype Reference Consortium and 1000 Genomes Project (1KGP), under UK Biobank application 27892. This work was conducted under a Not Human Subjects Research determination (NHSR-4298 at the Broad Institute of MIT and Harvard).

### UK Biobank phenotypes

Phenotype data processing and other downstream analyses were conducted using R version 3.6.0 [27]. HbA1c (UKB field 30750) was measured in blood samples through high-performance liquid chromatography (HPLC) using a Bio-Rad VARIANT II Turbo platform. HbA1c values and effect sizes are reported here in units of mmol/mol, as originally provided by UKB. Type 1 and type 2 diabetes were defined by expanding “probable” and “possible” definitions of diabetes from a previously developed algorithm [28] to take into account information from repeat assessment center visits and touchscreen diabetes diagnosis (UKB field 2443). After the removal of individuals with type 1 or type 2 diabetes, outliers for HbA1c were removed using a cutoff of 3 interquartile ranges below or above the 25^th^ and 75^th^ percentiles, respectively.

Dietary data processing was based on the procedure described by Cole and colleagues [12]. All dietary phenotype derivation was conducted on a homogenous population of unrelated individuals of European (EUR) ancestry (N=340,705), as determined by: 1) limiting to unrelated, high quality samples used in ancestry PCA conducted by UKB (Bycroft 2018), 2) projection of genotypes onto 1KGP phase 3 PCA space, 3) outlier detection to identify the largest cluster of individuals in which all clustered individuals fell within 1KGP EUR PC1 and PC2 limits, and 4) self-report as one of the following: “British”, “Irish”, “Any other white background”, “White”, “Do not know”, or “Prefer not to answer”.

The UKB FFQ consists of quantitative continuous variables (e.g. field 1289, tablespoons of cooked vegetables per day), ordinal non-quantitative variables depending on overall daily/weekly frequency (e.g. field 1329, overall oily fish intake), food types (i.e. milk, spread, bread, cereal, or coffee), or foods never eaten (field 6144, dairy, eggs, sugar, and wheat). Supplementary Table S2 provides a list of UKB fields relating to the corresponding FFQ question for each dietary habit, which can be viewed in the UKB Data Showcase (http://biobank.ndph.ox.ac.uk/showcase/). Ordinal variables were ranked and set to quantitative values, while food types or foods never eaten were converted into a series of binary variables. A single high-resolution overall alcohol intake variable was derived by combining questions on either “drinks per month” or “drinks per week” for each alcoholic beverage type (answered by different individuals depending on their reported overall alcohol frequency (field 1558)) while median-imputing values for those who reported consuming alcohol but responded either “Do not know” or “Prefer not to answer” to specific drink frequency.

Quantitative traits (QTs) were then inverse rank normal transformed and missing data were median-imputed for all traits. Dietary patterns (dietary principal components, or dPCs) represent the first principal component from a series of PCA (using the *prcomp* function in base R), including seven PCA on groups of related dietary traits and one global PCA using all 30 dietary traits (Supplementary Table S2). Given the substantial correlation between many of these traits, we derived an effective number of traits for multiple testing correction: PCA was conducted on the 38 scaled and mean-imputed traits, and the associated eigenvalues *λ* were used to calculate the number of effective traits as 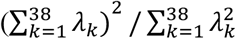 [29].

Additional phenotypes were retrieved for use in sensitivity models. Physical activity was calculated in units of excess metabolic equivalents (METs) as previously described [30], with median-imputation used to replace ambiguous values (“Prefer not to answer”, “Do not know”, or missing) for physical activity-related questionnaire items. Tobacco smoking behavior was assigned a score based on questionnaire responses: 0 (“No”), 1 (“Only occasionally”), or 2 (“Yes, on most or all days”). Educational attainment was derived in terms of US years of schooling equivalents as previously described [31], with median-imputation used to replace ambiguous values (“Prefer not to answer” or missing).

### Genome-wide interaction studies

For each dPC and individual dietary trait (38 exposures in total), a GWIS was performed using the following model:

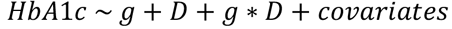

where g represents the imputed genotype dosage and D represents the dietary exposure. Covariates in the primary model included sex, age, age^2^, and 10 genetic principal components to control for population stratification estimated using FlashPCA2 [32]. Interaction analysis was performed using *GEM* (Gene-Environment interaction analysis in Millions of samples) [7] with robust standard error estimates.

An extensive set of sensitivity models were then applied to loci reaching genome-wide significance (*p* < 5×10^−8^). First, additional interaction parameters were included in the model between genotype and all covariates (sensitivity model 1 [SM1]) to adjust for possible confounding by the genotype × covariate interaction effect that is not accounted for by the covariate main effect [33]. Second, birthplace and study center (and their interactions with genotype) were included as additional covariates (SM2) to further account for potential coincident ancestry and cultural substructure in the population. Third, BMI, smoking, physical activity, and educational attainment (and their interactions with genotype) were additionally added to the model (SM3) to address confounding by specific socio-cultural factors. Fourth, SM3 was repeated while including individuals with diabetes (SM4) to test for the potential of collider bias via the diabetes exclusion criterion in the primary model.

Top variants were assigned to genes based on the NCBI dbSNP database where available, and otherwise based on the nearest gene within 10kb (in the case of the rs147678157-APOBEC3B assignment). PhenoScanner v2 [34,35] was used to explore variant associations with phenotypic traits and molecular quantities (e.g. eQTL as retrieved from the GTEx v7 release and methylation-quantitative trait loci [36]). Queries included proxy variants having r^2^ > 0.8 with the query variant in European populations. Stratified analyses were performed using linear regression models in R and the same set of covariates as in the primary interaction analysis. Separate stratified models were performed with respect to the relevant dietary trait (quintiles) and genotype (three groups, with hard-calls generated by rounding dosages to the nearest genotype [0/1/2]). Stratified analyses were performed in two ways: testing for the effect of hard-called genotypes among each of 5 dietary trait quintiles, and vice versa, testing for the effect of the dietary trait in each of 3 genotype groups. Stratified models were tested using base linear regression models in R with the same set of covariates as in the primary interaction analysis.

### Multi-ancestry replication and testing in related glycemic traits

Multi-ancestry replication was performed using three non-European ancestry groups in UKB (West African, East Asian, and South Asian). Using 1KGP phase 3 as the training dataset, we built probabilistic Gaussian mixture models to represent normally-distributed subpopulations within the overall population, assigning data points to the multivariate normal components that maximize the component posterior probability. Ten-fold cross-validation was used with different initialization states and clustering was evaluated using the adjusted Rand index score. The model with the highest adjusted Rand index score for each ancestry was used to cluster individuals in UKB into ancestry subgroups based on their 1KGP projected PCs and self-reported ethnicities (field 2100). Multi-ancestry dietary and clinical phenotypes were derived as described above for European individuals, and dPCs were derived by projecting ancestry-specific dietary traits onto the European principal component loadings. Interaction analysis was conducted for top variant-trait pairs under the primary model described above in Europeans.

To test the robustness of the top associations, two GDI analyses using related glycemic traits (follow-up HbA1c and fasting glucose levels) were performed based on the primary model in Europeans. For the follow-up analysis, all variables were taken from the second assessment center visit (i.e. the first follow-up), with dPCs projected as described above in the multi-ancestry analysis. For the fasting glucose analysis, all phenotypes and analysis parameters were identical to the primary model in Europeans, other than the replacement of HbA1c as the outcome with fasting glucose, defined as random glucose in participants who reported having fasted for at least 8 hours prior to the blood draw (UKB fields 30740 and 74).

General agreement between the primary results and replication results was tested via an enrichment test for shared interaction effect signs. Suggestive variant-trait pairs, defined as having interaction *p* < 1×10^−5^ in the primary analysis, were collected and pruned using 500kb windows. For each within-UKB replication approach, a Chi-square test for shared signs of interaction effects was performed.

Replication was also undertaken in the Women’s Genome Health Study, a prospective US-based cohort of ∼23,000 females 45 years or older, described in detail elsewhere [37]. After quality control procedures, the final interaction analysis was conducted in 20,095 females of European ancestry, free of a history of diabetes, and whose current diet had not “greatly” changed in comparison to the last 5 years. The HbA1c phenotype was processed in a similar manner as described for UK Biobank. Semi-quantitative dietary exposures and fruit and vegetable food groups were converted to continuous variables as described previously [38] and inverse normal transformed. Using an Rsq cutoff of 0.50, three variant-trait pairs were tested for replication. The rs140270534 (dried fruit) interaction was tested with overall fruit intake in WGHS, the rs3010439 (fruit and vegetable dPC) was tested with overall fruit and vegetable intake in WGHS, and the 4:131148078 indel (milk) was tested with milk consumption in WGHS derived by binarizing individuals who drink any amount of skim or whole milk versus those that answer never or rarely for both. Effect sizes on HbA1c were converted from % to mmol/mol.

## Data Availability

UK Biobank data can be accessed at https://www.ukbiobank.ac.uk/.

## AUTHOR CONTRIBUTIONS

KEW – Conceptualization, Data Curation, Formal analysis, Software, Writing – Original Draft Preparation

JM – Data Curation, Software

DIC – Validation, Writing – Review &Editing

JCF – Supervision, Writing – Review &Editing

HC – Methodology, Software

AKM – Conceptualization, Funding Acquisition, Supervision, Writing – Review &Editing

JBC – Conceptualization, Data Curation, Supervision, Writing – Original Draft Preparation

## NOTES/ACKNOWLEDGEMENTS

While KEW and AKM are employees of Mass General Brigham (MGB), this work was not conducted in their capacity as MGB employees.

## REFERENCES

1. Zeevi D, Korem T, Zmora N, Israeli D, Rothschild D, Weinberger A, et al. Personalized Nutrition by Prediction of Glycemic Responses. Cell. 2015;163: 1079–1094. doi:10.1016/j.cell.2015.11.001

2. Barrington WT, Wulfridge P, Wells AE, Rojas CM, Howe SYF, Perry A, et al. Improving Metabolic Health Through Precision Dietetics in Mice. Genetics. 2018;208: 399–417. doi:10.1534/genetics.117.300536

3. Zheng J-S, Arnett DK, Lee Y-C, Shen J, Parnell LD, Smith CE, et al. Genome-Wide Contribution of Genotype by Environment Interaction to Variation of Diabetes-Related Traits. PLoS One. 2013;8: e77442. doi:10.1371/journal.pone.0077442

4. Franks PW, Merino J. Gene-lifestyle interplay in type 2 diabetes. Curr Opin Genet Dev. 2018;50: 35–40. doi:10.1016/j.gde.2018.02.001

5. Thomas D. Methods for Investigating Gene-Environment Interactions in Candidate Pathway and Genome-Wide Association Studies. Annu Rev Public Health. 2010;31: 21–36. doi:10.1146/annurev.publhealth.012809.103619

6. Gauderman WJ, Mukherjee B, Aschard H, Hsu L, Lewinger JP, Patel CJ, et al. Update on the State of the Science for Analytical Methods for Gene-Environment Interactions. Am J Epidemiol. 2017;186: 762–770. doi:10.1093/aje/kwx228

7. Westerman KE, Pham DT, Hong L, Chen Y, Sevilla-González M, Sung YJ, et al. Scalable and flexible gene-environment interaction analysis in millions of samples. bioRxiv. 2020; doi: https://doi.org/10.1101/2020.05.13.090803. doi:10.1101/2020.05.13.090803

8. Bi W, Zhao Z, Dey R, Fritsche LG, Mukherjee B, Lee S. A Fast and Accurate Method for Genome-wide Scale Phenome-wide G × E Analysis and Its Application to UK Biobank. Am J Hum Genet. 2019;105: 1182–1192. doi:10.1016/j.ajhg.2019.10.008

9. Tyrrell J, Wood AR, Ames RM, Yaghootkar H, Beaumont RN, Jones SE, et al. Gene–obesogenic environment interactions in the UK Biobank study. Int J Epidemiol. 2017;46: 559–575. doi:10.1093/ije/dyw337

10. Moore R, Casale FP, Jan Bonder M, Horta D, Franke L, Barroso I, et al. A linear mixed-model approach to study multivariate gene–environment interactions. Nat Genet. 2019;51: 180–186. doi:10.1038/s41588-018-0271-0

11. Tucker KL, Smith CE, Lai C-Q, Ordovas JM. Quantifying Diet for Nutrigenomic Studies. Annu Rev Nutr. 2013;33: 349–371. doi:10.1146/annurev-nutr-072610-145203

12. Cole JB, Florez JC, Hirschhorn JN. Comprehensive genomic analysis of dietary habits in UK Biobank identifies hundreds of genetic associations. Nat Commun. 2020;11: 1467. doi:10.1038/s41467-020-15193-0

13. MacDonald PE. TRP-ing Down the Path to Insulin Secretion. Diabetes. 2011;60: 28–29. doi:10.2337/db10-1402

14. Võsa U, Claringbould A, Westra H-J, Bonder MJ, Deelen P, Zeng B, et al. Unraveling the polygenic architecture of complex traits using blood eQTL meta-analysis. bioRxiv. 2018. doi:10.1101/447367

15. Ketema EB, Kibret KT. Correlation of fasting and postprandial plasma glucose with HbA1c in assessing glycemic control; systematic review and meta-analysis. Arch Public Heal. 2015;73: 43. doi:10.1186/s13690-015-0088-6

16. Vriens J, Nilius B, Vennekens R. Herbal compounds and toxins modulating TRP channels. Curr Neuropharmacol. 2008.

17. Uchida K, Tominaga M. TRPM2 modulates insulin secretion in pancreatic β-cells. Islets. 2011;3: 209–211. doi:10.4161/isl.3.4.16130

18. Wagner TFJ, Loch S, Lambert S, Straub I, Mannebach S, Mathar I, et al. Transient receptor potential M3 channels are ionotropic steroid receptors in pancreatic β cells. Nat Cell Biol. 2008. doi:10.1038/ncb1801

19. Straub I, Mohr F, Stab J, Konrad M, Philipp S, Oberwinkler J, et al. Citrus fruit and fabacea secondary metabolites potently and selectively block TRPM3. Br J Pharmacol. 2013;168: 1835– 1850. doi:10.1111/bph.12076

20. Palmer C, Pe’er I. Statistical correction of the Winner’s Curse explains replication variability in quantitative trait genome-wide association studies. Marchini J, editor. PLOS Genet. 2017;13: e1006916. doi:10.1371/journal.pgen.1006916

21. Aschard H. A perspective on interaction effects in genetic association studies. Genet Epidemiol. 2016;40: 678–688. doi:10.1002/gepi.21989

22. Li SX, Imamura F, Ye Z, Schulze MB, Zheng J, Ardanaz E, et al. Interaction between genes and macronutrient intake on the risk of developing type 2 diabetes: systematic review and findings from European Prospective Investigation into Cancer (EPIC)-InterAct. Am J Clin Nutr. 2017;106: 263–275. doi:10.3945/ajcn.116.150094

23. Kraft P, Aschard H. Finding the missing gene–environment interactions. Eur J Epidemiol. 2015. doi:10.1007/s10654-015-0046-1

24. Willett WC, Howe GR, Kushi LH. Adjustment for total energy intake in epidemiologic studies. Am J Clin Nutr. 1997;65: 1220S–1228S. doi:10.1093/ajcn/65.4.1220S

25. Sudlow C, Gallacher J, Allen N, Beral V, Burton P, Danesh J, et al. UK Biobank: An Open Access Resource for Identifying the Causes of a Wide Range of Complex Diseases of Middle and Old Age. PLOS Med. 2015;12: e1001779. doi:10.1371/journal.pmed.1001779

26. Bycroft C, Freeman C, Petkova D, Band G, Elliott LT, Sharp K, et al. The UK Biobank resource with deep phenotyping and genomic data. Nature. 2018;562: 203–209. doi:10.1038/s41586-018-0579-z

27. R Core Team. R: A Language and Environment for Statistical Computing. Vienna, Austria: R Foundation for Statistical Computing; 2019. Available: https://www.r-project.org/

28. Eastwood S V., Mathur R, Atkinson M, Brophy S, Sudlow C, Flaig R, et al. Algorithms for the Capture and Adjudication of Prevalent and Incident Diabetes in UK Biobank. Herder C, editor. PLoS One. 2016;11: e0162388. doi:10.1371/journal.pone.0162388

29. Bretherton CS, Widmann M, Dymnikov VP, Wallace JM, Bladé I. The Effective Number of Spatial Degrees of Freedom of a Time-Varying Field. J Clim. 1999;12: 1990–2009. doi:10.1175/1520-0442(1999)012<1990:TENOSD>2.0.CO;2

30. Guo W, Bradbury KE, Reeves GK, Key TJ. Physical activity in relation to body size and composition in women in UK Biobank. Ann Epidemiol. 2015;25: 406-413.e6. doi:10.1016/j.annepidem.2015.01.015

31. Ge T, Chen C-Y, Doyle AE, Vettermann R, Tuominen LJ, Holt DJ, et al. The Shared Genetic Basis of Educational Attainment and Cerebral Cortical Morphology. Cereb Cortex. 2019;29: 3471–3481. doi:10.1093/cercor/bhy216

32. Abraham G, Qiu Y, Inouye M. FlashPCA2: principal component analysis of Biobank-scale genotype datasets. Stegle O, editor. Bioinformatics. 2017;33: 2776–2778. doi:10.1093/bioinformatics/btx299

33. Keller MC. Gene × Environment Interaction Studies Have Not Properly Controlled for Potential Confounders: The Problem and the (Simple) Solution. Biol Psychiatry. 2014;75: 18–24. doi:10.1016/j.biopsych.2013.09.006

34. Kamat MA, Blackshaw JA, Young R, Surendran P, Burgess S, Danesh J, et al. PhenoScanner V2: an expanded tool for searching human genotype–phenotype associations. Kelso J, editor. Bioinformatics. 2019;35: 4851–4853. doi:10.1093/bioinformatics/btz469

35. Staley JR, Blackshaw J, Kamat MA, Ellis S, Surendran P, Sun BB, et al. PhenoScanner: a database of human genotype–phenotype associations. Bioinformatics. 2016;32: 3207–3209. doi:10.1093/bioinformatics/btw373

36. Bonder MJ, Luijk R, Zhernakova D V., Moed M, Deelen P, Vermaat M, et al. Disease variants alter transcription factor levels and methylation of their binding sites. Nat Genet. 2017. doi:10.1038/ng.3721

37. Ridker PM, Chasman DI, Zee RYL, Parker A, Rose L, Cook NR, et al. Rationale, Design, and Methodology of the Women’s Genome Health Study: A Genome-Wide Association Study of More Than 25 000 Initially Healthy American Women. Clin Chem. 2008;54: 249–255. doi:10.1373/clinchem.2007.099366

38. Liu S, Manson JE, Lee I-M, Cole SR, Hennekens CH, Willett WC, et al. Fruit and vegetable intake and risk of cardiovascular disease: the Women’s Health Study. Am J Clin Nutr. 2000;72: 922–928. doi:10.1093/ajcn/72.4.922

