## Supplementary Materials for "Genome-wide gene-diet interaction analysis in the UK Biobank identifies novel effects on Hemoglobin A1c"

### Supplementary Figures

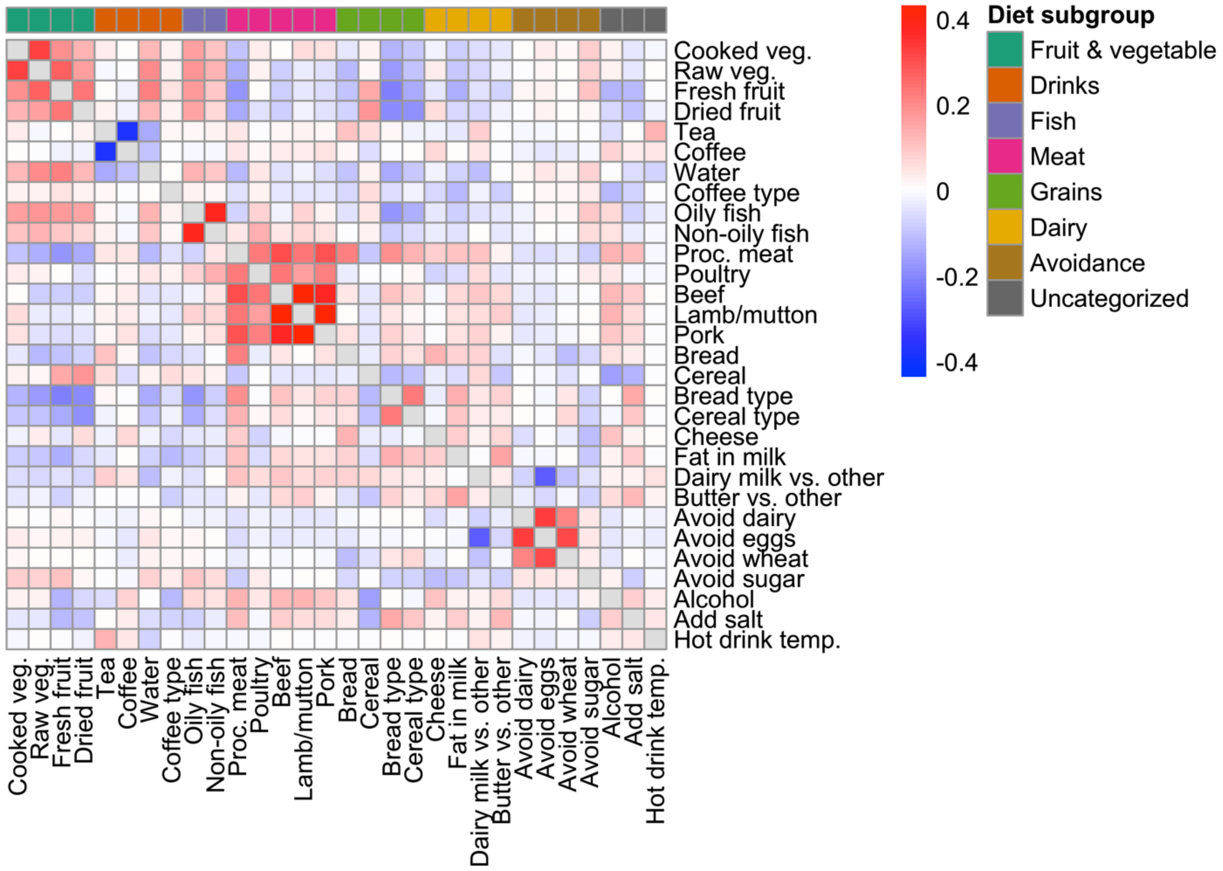


**Supplementary Figure S1:** Correlations between dietary traits. Heatmap shows Pearson correlations between individual dietary traits. Colors in the top bar correspond to the assigned dietary trait subgroup.

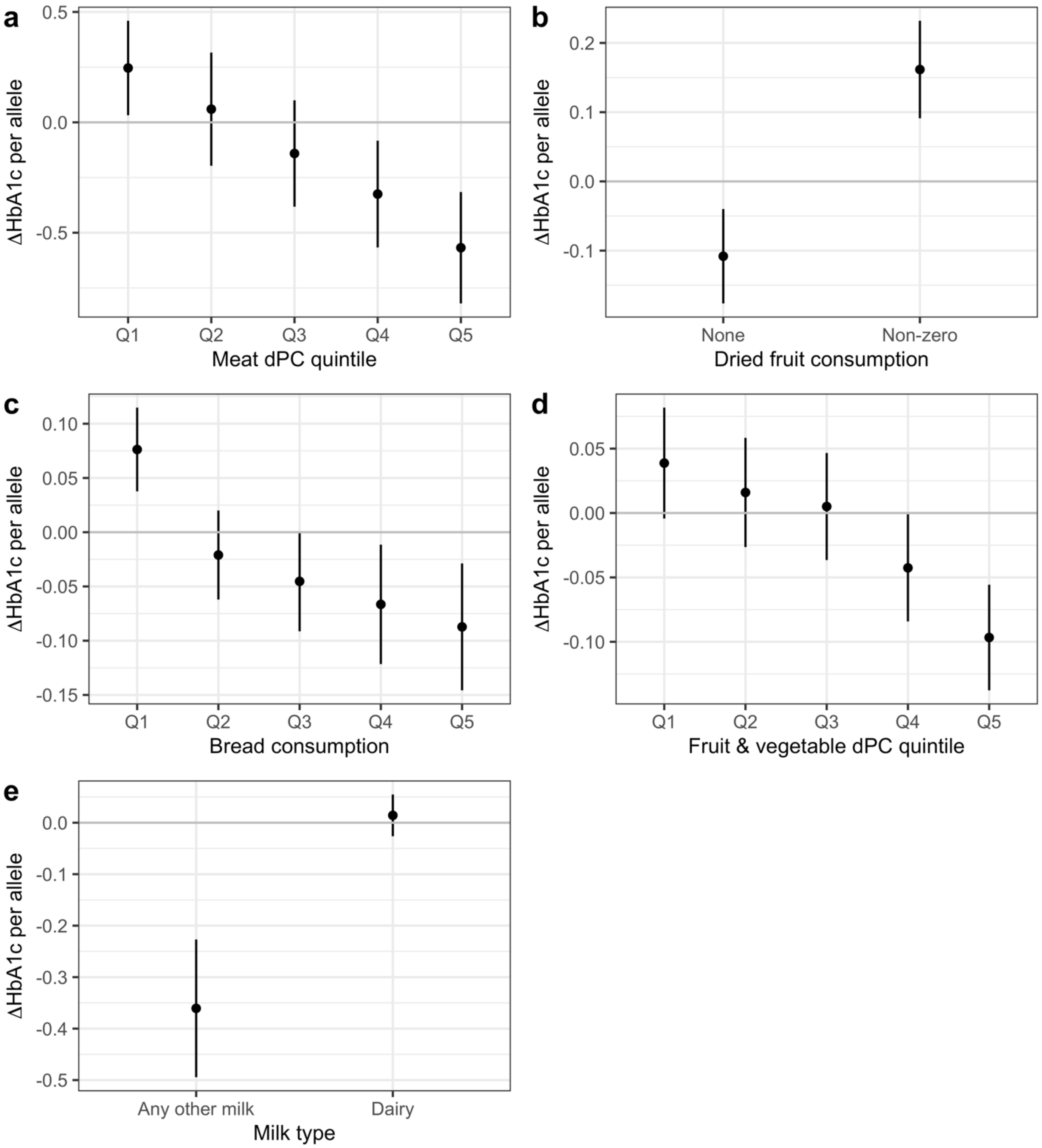


**Supplementary Figure S2:** Diet-stratified effects at genome-wide significant index variants. a) rs147678157 genotype effect sizes on HbA1c (y-axis) after stratification into quintiles of meat dPC values (x-axis). b) As in (a), but instead using rs140270534 genotypes and dried fruit consumption. c) As in (a), but instead using genotypes at rs62218803 and bread consumption. d) As in (a), but instead using rs3010439 genotypes and fruit & vegetable dPC. e) As in (a), but instead using rs3010439 genotypes and milk type. Points and bars denote stratified main effect estimates and 95% confidence intervals from the primary model. Genotype-stratified effects are shown only for genotype groups containing >100 individuals. Effect estimates are in terms of mmol/mol HbA1c. s.d.: standard deviation.

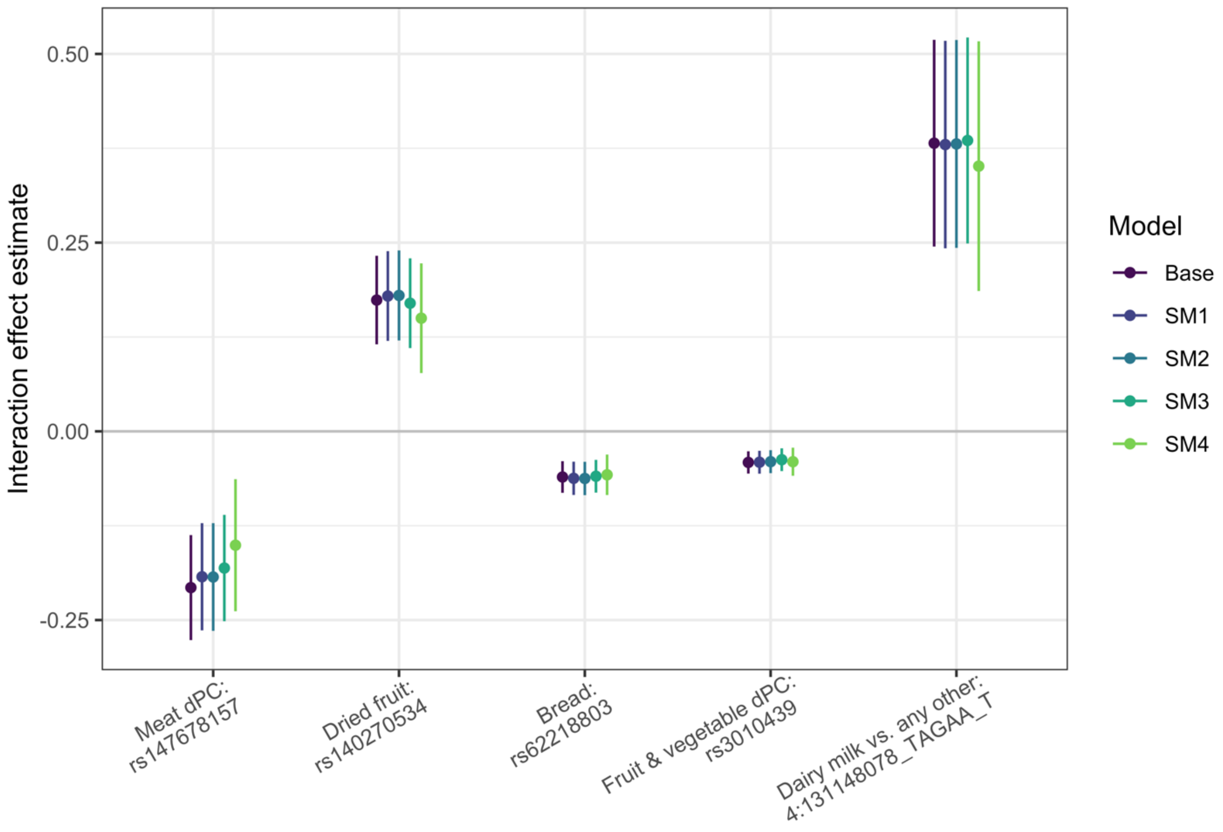


**Supplementary Figure S3:** Results from sensitivity models at genome-wide significant ($p < 5x10^{-8}$ index variants from the primary analysis. Colors correspond to sensitivity models (SM1: inclusion of genotype-by-covariate interactions for all covariates, SM2: additional adjustment for birthplace and assessment center, SM3: additional adjustment for BMI, smoking, educational attainment, and physical activity), SM4: as in SM3, but including individuals with diabetes). Error bars denote 95% confidence intervals for the interaction effect estimates.

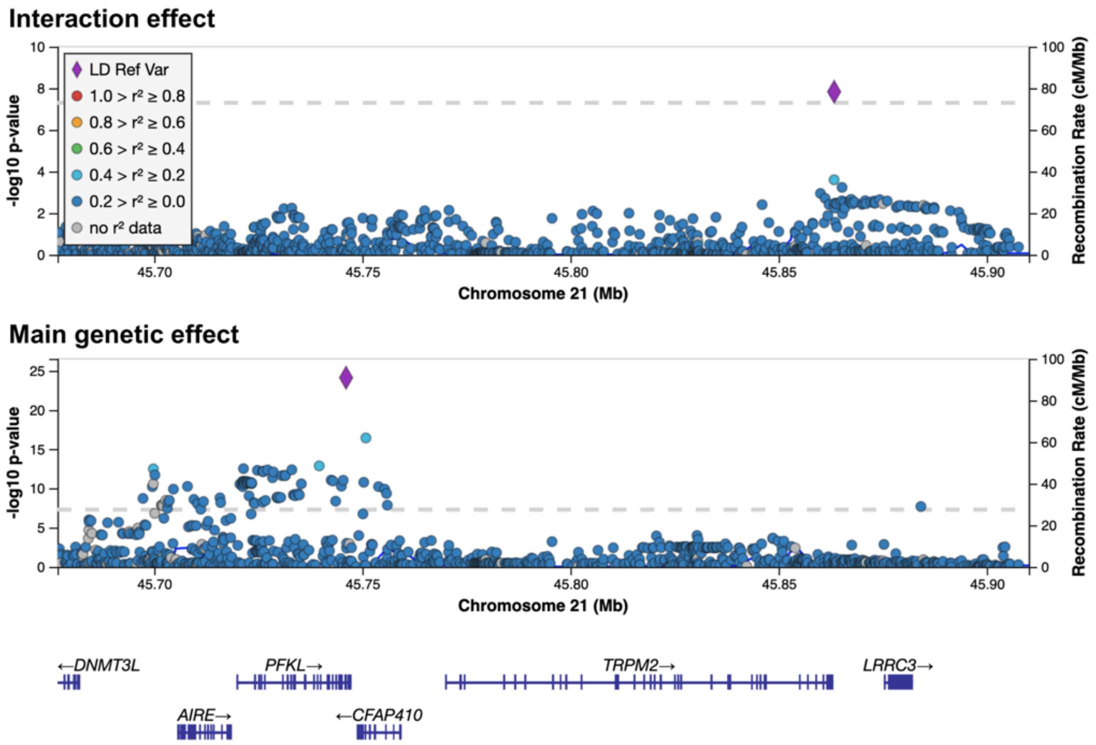


**Supplementary Figure S4:** Regional plot shows minimum -$\log_{10}$ ($p$-values) for the bread consumption interaction effect (top panel) and genetic main effect (bottom panel) as a function of genomic position. Colors correspond to linkage disequilibrium with the reference variant (purple) in each panel. s.d.: standard deviation.

### Supplementary Tables

**Supplementary Table S1:** UK Biobank population description.

| Trait | Value |
|---|---|
| **N** | 340705 |
| **Sex** | 50% female |
| **Age in years** | 57 (8) |
| **Current smoking** | 10.4% |
| **BMI** | 27.1 (4.6) |
| **HbA1c (mmol/mol)** | 35 (3.7) |
| **Physical activity in METs/wk** | 48.1 (62) |
| **Educational attainment in US school yr. equivalents** | 15 (5) |

Continuous variables are represented as: mean (standard deviation). BMI: body mass index, METs: metabolic equivalents.

**Supplementary Table S2:** Derived dietary traits used in this study.

| Dietary trait | UKB Source Field | Food sub-group |
|---|---|---|
| **Cooked vegetables** | 1289 | Fruit & vegetable |
| **Raw vegetables** | 1299 | Fruit & vegetable |
| **Fresh fruit** | 1309 | Fruit & vegetable |
| **Dried fruit** | 1319 | Fruit & vegetable |
| **Tea** | 1488 | Drinks |
| **Coffee** | 1498 | Drinks |
| **Water** | 1528 | Drinks |
| **Decaf vs. regular coffee** | 1508 | Drinks |
| **Oily fish** | 1329 | Fish |
| **Non-oily fish** | 1339 | Fish |
| **Processed meat** | 1349 | Meat |
| **Poultry** | 1359 | Meat |
| **Beef** | 1369 | Meat |
| **Lamb/mutton** | 1379 | Meat |
| **Pork** | 1389 | Meat |
| **Bread** | 1438 | Grains |
| **Cereal** | 1458 | Grains |
| **White bread vs. brown or whole grain** | 1448 | Grains |
| **Whole grain cereal vs. other types** | 1468 | Grains |
| **Cheese** | 1408 | Dairy |
| **Level of fat in milk** | 1418 | Dairy |
| **Dairy milk vs. any other** | 1418 | Dairy |
| **Butter vs. other spreads** | 1428 | Dairy |
| **Avoid dairy** | 6144 | Avoidance |
| **Avoid eggs** | 6144 | Avoidance |
| **Avoid wheat** | 6144 | Avoidance |
| **Avoid sugar** | 6144 | Avoidance |
| **Alcohol** | 1558, 1568, 1578, 1588, 1598, 1608, 4407, 4418, 4429, 4440, 4451, 4462, 5364 | |
| **Add salt to food** | 1478 | |
| **Hot drink temperature** | 1518 | |

**Supplementary Table S3:** Associations of genome-wide significant index variants with alternate dietary traits and dPCs.

| Dietary trait | P - rs3010439 | P - rs140270534 | P - rs147678157 | P - rs62218803 | P - 4:131148078_TAGAA_T |
|---|---|---|---|---|---|
| **Fruit & vegetable dPC** | 4.53E-08 | 6.87E-05 | >0.01 | >0.01 | >0.01 |
| **Meat dPC** | >0.01 | >0.01 | 5.45E-09 | >0.01 | >0.01 |
| **Grains dPC** | >0.01 | 0.0035 | >0.01 | 0.00959 | >0.01 |
| **Global dPC** | >0.01 | 0.00245 | 9.84E-06 | >0.01 | >0.01 |
| **Cooked vegetables** | 9.84E-07 | 0.0043 | >0.01 | >0.01 | >0.01 |
| **Raw vegetables** | 0.00166 | >0.01 | >0.01 | >0.01 | >0.01 |
| **Fresh fruit** | 0.000423 | >0.01 | >0.01 | >0.01 | >0.01 |
| **Dried fruit** | 0.00643 | 6.25E-09 | >0.01 | >0.01 | >0.01 |
| **Bread** | >0.01 | >0.01 | >0.01 | 1.47E-08 | >0.01 |
| **Tea** | >0.01 | >0.01 | 0.00589 | >0.01 | >0.01 |
| **Processed meat** | >0.01 | >0.01 | 0.000201 | >0.01 | >0.01 |
| **Beef** | >0.01 | >0.01 | 3.72E-05 | >0.01 | >0.01 |
| **Lamb/mutton** | >0.01 | >0.01 | 2.98E-05 | >0.01 | >0.01 |
| **Pork** | >0.01 | >0.01 | 1.68E-06 | >0.01 | >0.01 |
| **Avoid wheat** | >0.01 | >0.01 | >0.01 | 0.00709 | >0.01 |
| **Dairy milk vs. any other** | >0.01 | >0.01 | >0.01 | >0.01 | 4.66E-08 |
| **White bread vs. brown/WG** | >0.01 | 0.00907 | >0.01 | >0.01 | >0.01 |

All numeric cells represent interaction *p*-values from the primary model. Diet traits and dPCs not shown did not interact at $p < 0.01$ with any of the variants in this table.

**Supplementary Table S4:** Results from sensitivity models at genome-wide significant variant-trait pairs from the primary analysis.

| Dietary trait | Index variant | Diet effect | Genetic main effect | Interaction effect - Base | Interaction effect - SM1 | Interaction effect - SM2 | Interaction effect - SM3 | Interaction effect - SM4 |
|---|---|---|---|---|---|---|---|---|
| **Meat dPC** | rs14767815 7 | 0.14 (7.22x10$^{-115}$ | -0.131 (0.0156) | -0.207 (5.45x10$^{-09}$ | -0.193 (1.05x10$^{-07}$ | -0.193 (1.14x10$^{-07}$ | -0.181 (4.69x10$^{-07}$ | -0.151 (0.000724) |
| **Dried fruit** | rs14027053 4 | -0.141 (5.08x10$^{-115}$ | 0.0155 (0.534) | 0.174 (6.25x10$^{-09}$ | 0.179 (3.43x10$^{-09}$ | 0.18 (3.31x10$^{-09}$ | 0.17 (2.2x10$^{-08}$ | 0.15 (5.28x10$^{-05}$ |
| **Bread** | rs62218803 | 0.18 (2.08x10$^{-173}$ | -0.0138 (0.19) | -0.0605 (1.47x10$^{-08}$ | -0.0622 (3.11x10$^{-08}$ | -0.0625 (3.08x10$^{-08}$ | -0.0595 (7.64x10$^{-08}$ | -0.0576 (2.55x10$^{-05}$ |
| **Fruit & vegetable dPC** | rs3010439 | -0.189 (4.72x10$^{-202}$ | -0.0155 (0.106) | -0.0412 (4.53x10$^{-08}$ | -0.0409 (1.19x10$^{-07}$ | -0.0402 (2.21x10$^{-07}$ | -0.0376 (1.08x10$^{-06}$ | -0.0402 (2.15x10$^{-05}$ |
| **Dairy milk vs. any other** | 4:13114807 8_TAGAA_ T | 0.0785 (2.44x10$^{-38}$ | -0.0168 (0.396) | 0.382 (4.66x10$^{-08}$ | 0.38 (6.08x10$^{-08}$ | 0.381 (5.97x10$^{-08}$ | 0.385 (3.11x10$^{-08}$ | 0.351 (3.14x10$^{-05}$ |

Effects are shown as: coefficient (*p*-value).
[1]Sensitivity model 1 includes genotype interaction terms for all covariates
[2]Sensitivity model 2 additionally includes indicator variables for assessment center and birthplace (and interactions)
[3]Sensitivity model 3 additionally includes BMI, smoking, educational attainment, and physical activity (and interactions)
[4]Sensitivity model 4 additionally includes individuals with type 1 and type 2 diabetes

**Supplementary Table S5:** Multi-ancestry replication of genome-wide significant index variants associated with dPCs at the constituent trait level.

| Dietary trait | Index variant | EUR Effect (P) | AFR Effect (P) | EAS Effect (P) | SAS Effect (P) |
|---|---|---|---|---|---|
| **Fruit & vegetable dPC** | rs3010439 | 0.041 (4.5x10$^{-08}$ | -0.102 (0.22) | 0.117 (0.08) | -0.058 (0.59) |
| **Fresh fruit** | rs3010439 | | 0.306 (0.0082) | -0.01 (0.92) | -0.008 (0.95) |
| **Cooked vegetables** | rs3010439 | | 0.031 (0.8) | -0.239 (0.012) | 0.238 (0.082) |
| **Raw vegetables** | rs3010439 | | -0.103 (0.37) | -0.115 (0.23) | 0.029 (0.83) |
| **Dried fruit** | rs3010439 | | 0.049 (0.72) | -0.101 (0.31) | 0.117 (0.44) |
| **Meat dPC** | rs147678157 | -0.207 (5.5x10$^{-09}$ | -1.625 (0.2) | 1.471 (0.091) | 1.332 (0.72) |
| **Pork** | rs147678157 | | 1.112 (0.5) | 2.49 (0.1) | 3.374 (0.39) |
| **Processed meat** | rs147678157 | | -3.609 (0.47) | 3.239 (0.022) | 8.369 (0.21) |
| **Lamb/mutton** | rs147678157 | | -0.221 (0.89) | 2.035 (0.079) | -3.789 (0.33) |
| **Beef** | rs147678157 | | -2.237 (0.35) | 2.044 (0.15) | -1.995 (0.6) |
| **Poultry** | rs147678157 | | -2.282 (0.27) | -0.38 (0.55) | -1.062 (0.73) |

**Supplementary Table S6:** Within-population validation of genome-wide significant index variants.

| Diet trait | Index variant | Effect-Base | Effect-FU | Effect-FG |
|---|---|---|---|---|
| **Meat dPC** | rs147678157 | -0.207 (5.5x$10^{-09}$ | -0.219 (0.24) | -0.053 (0.02) |
| **Dried fruit** | rs140270534 | 0.174 (6.3x$10^{-09}$ | 0.114 (0.47) | 0.031 (0.23) |
| **Bread** | rs62218803 | -0.061 (1.5x$10^{-08}$ | -0.1 (0.072) | -0.004 (0.59) |
| **Fruit and vegetable dPC** | rs3010439 | 0.041 (4.5x$10^{-08}$ | -0.059 (0.16) | -0.007 (0.2) |
| **Dairy milk vs. any other** | 4:131148078_TAGAA_T | 0.382 (4.7x$10^{-08}$ | 1.083 (0.0047) | 0.063 (0.22) |

Cells display beta (*p*-value) for interaction effect estimate. FU: follow-up, FG: fasting glucose.

**Supplementary Table S7:** Replication of genome-wide significant index variants in the Women's Genome Health Study dataset.

| Dietary trait | Variant | Effect allele | Effect allele frequency | Genetic main effect | Interaction effect |
|---|---|---|---|---|---|
| **Fruit** | rs140270534 | G | 0.046 | 0.0751 (0.363) | -0.045 (0.591) |
| **Fruits & vegetables** | rs3010439 | T | 0.718 | -0.00652 (0.837) | 0.0138 (0.663) |
| **Dairy milk vs. none** | 4:131148078_TAGAA_T | T | 0.053 | 0.191 (0.323) | -0.239 (0.245) |

Effects are shown as: coefficient (*p*-value). Replication model in WGHS contains the same covariates as the primary model, other than sex and genotyping array (age, age-squared, and ten genetic PCs).
